# Optimizing SARS-CoV-2 Variant of Concern Screening: Experience from British Columbia, Canada, Early 2021

**DOI:** 10.1101/2021.03.23.21253520

**Authors:** Catherine A. Hogan, Hind Sbihi, Agatha Jassem, Natalie Prystajecky, John R. Tyson, Tracy D. Lee, Frankie Tsang, Corrinne Ng, Loretta Janz, Marc G. Romney, Linda Hoang

**Affiliations:** British Columbia Centre for Disease Control Public Health Laboratory, Vancouver, British Columbia, Canada; School of Population and Public Health, University of British Columbia, Vancouver, British Columbia, Canada; Department of Pathology and Laboratory Medicine, University of British Columbia, Vancouver, British Columbia, Canada; Division of Medical Microbiology and Virology, St. Paul’s Hospital, Vancouver, British Columbia, Canada

**Keywords:** SARS-CoV-2, variant of concern, testing, public health, COVID-19

## Abstract

Comprehensive and timely testing is required for SARS-CoV-2 variant of concern (VoC) screening. Whole genome sequencing (WGS) provides the broadest means to detect circulating VoCs, but requires longer turnaround time than targeted molecular testing by quantitative polymerase chain reaction (qPCR). We demonstrated the feasibility of a combined testing approach for VoC prevalence assessment in British Columbia, and showed high concordance between qPCR testing and WGS. This directly informed wider VoC screening strategy implementation, and public health efforts.

## Introduction

Identification of mutations in the SARS-CoV-2 genome has led to the recognition of variants of concern (VoCs) based on their association with increased transmissibility, clinical severity, and/or reduced vaccine efficacy (1-5). Globally, lineages B.1.1.7, B.1.351, and P.1 represent the three currently circulating VoCs (6). Whole genome sequencing (WGS) provides the most comprehensive means to detect circulating VoCs, but requires considerable resources and time to perform. VoC screening workflows using quantitative polymerase chain reaction (qPCR) optimize testing capacity and turnaround time to enable appropriate and timely public health interventions (7, 8).

## Methods

The British Columbia Centre for Disease Control Public Health Laboratory (BCCDC PHL) serves as the reference laboratory for the province. A combined VoC testing strategy based on direct WGS and targeted N501Y mutation qPCR was employed to estimate point prevalence (**Appendix Figure 1**) based on samples resulted as positive for SARS-CoV-2 from January 30^th^ to February 5^th^ 2021. Specimens not tested directly by WGS were screened by an assay targeting the envelope (*E*) gene and N501Y mutation, which detects but does not differentiate the three main VoCs, in two laboratories (**Supplementary methods**). Specimens with a detected N501Y mutation underwent confirmatory testing and typing by WGS (**Supplementary methods**).

Ongoing surveillance was continued from February 6^th^ to March 6^th^ 2021 to monitor evolution in VoC prevalence. Epidemiologic and laboratory data collection was performed based on linkage of provincial databases. This work was conducted under the public health mandate and institutional review board approval was waived.

## Results

A total of 3,024 specimens underwent either direct WGS or screening by N501Y assay, representing over 97% of the 3,101 SARS-CoV-2-positive samples in British Columbia for the sampling period (**Figure 1**). The prevalence of VoCs by both pathways, direct WGS and N501Y qPCR screening, was below 1%. Of the 27 individuals with a confirmed VoC, the median age was 28 years (interquartile range, 20-40), and 11 were female. A total of 23 B.1.1.7 and 4 B.1.351 lineages were confirmed, and there was no P.1 lineage identified during this period (**Table 1**). Geographical stratification revealed confirmed cases were concentrated in two health authorities (**Appendix Figure 2)**. Furthermore, direct WGS enabled the identification of one case of B.1.525 lineage (**Appendix Figure 2**), which pointed to the recent emergence of a lineage of Nigerian origin in Canada during this time period (9). Continued surveillance by N501Y screening revealed a progressive increase in VoC prevalence in British Columbia, reaching over 10% by the end of February 2021 (**Appendix Table 1**).

**Figure 1.**
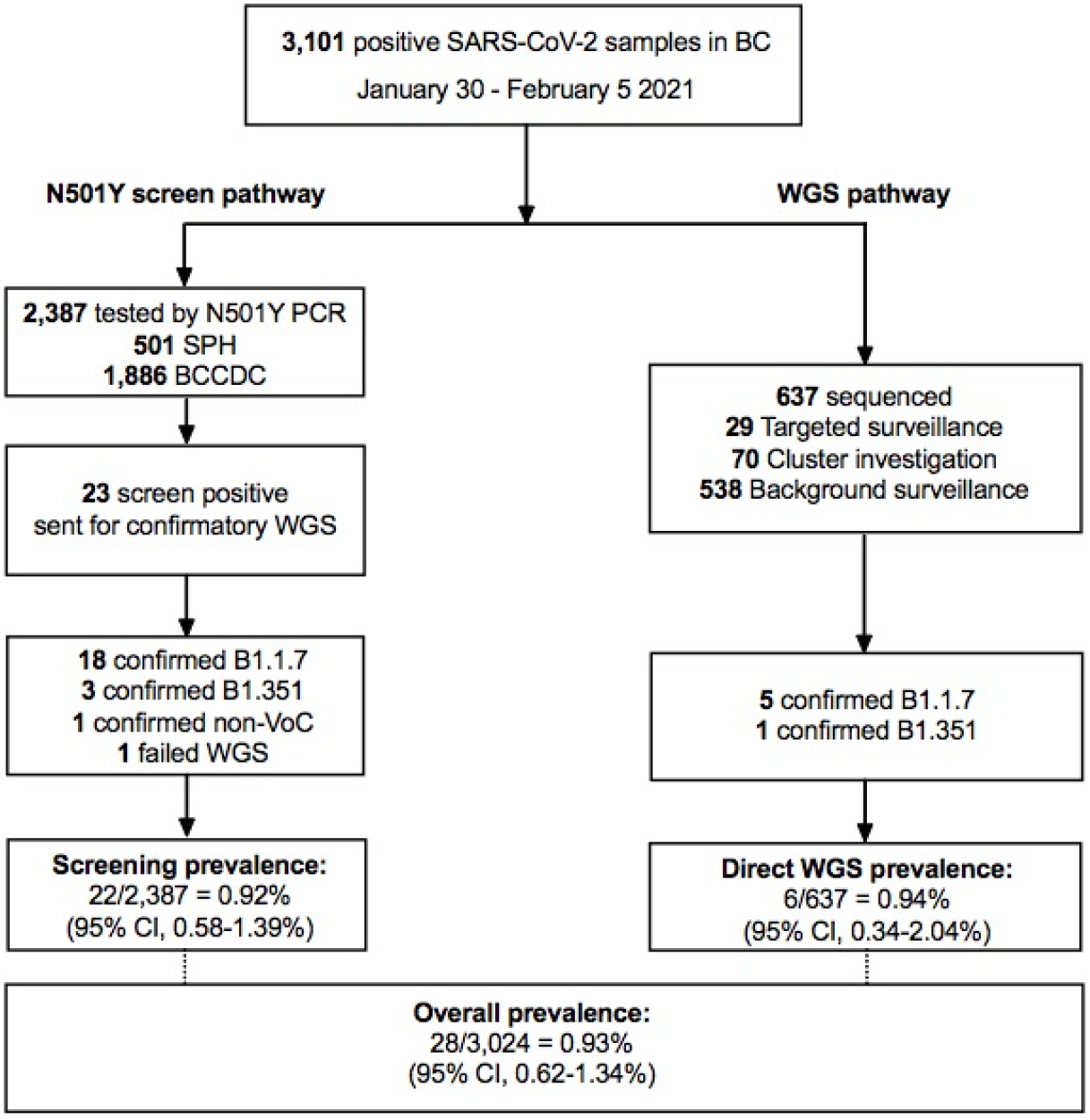
Flowchart of the screening and direct WGS testing results. The prevalence of SARS-CoV-2 variants of concern was <1% in both pathways. Most specimens were identified as B.1.1.7 lineage, and there was no P.1 lineage identified. One specimen underwent WGS twice but generated only an incomplete genome; for prevalence analysis, this specimen was considered as a confirmed VoC. BC: British Columbia; BCCDC: British Columbia Centre for Disease Control; CI: confidence interval; PCR: polymerase chain reaction; SARS-CoV-2: severe acute respiratory syndrome coronavirus 2; SPH: St Paul’s Hospital; VoC: variant of concern; WGS: whole genome sequencing.

**Table 1.** Epidemiologic and laboratory characteristics of the 28 individuals with presumptive or confirmed SARS-CoV-2 variants of concern identified in the study.

| Detection pathway | Subject | Age group/<br>sex | Sample type | *E* gene<br>Ct<br>result | WGS<br>indication | WGS<br>result | Lineage |
| --- | --- | --- | --- | --- | --- | --- | --- |
| VoC<br>screening | VoC-1 | 18-50yo; F | NP | 22.47 | N501Y detected | Failed | NA |
|  | VoC-2 | 18-50yo; M | SG | 19.28 | N501Y detected | Yes | B.1.1.7 |
|  | VoC-3 | >50yo; M | NP | NA | N501Y detected | Yes | B.1.351 |
|  | VoC-4 | >50yo; M | NA | 25.78 | N501Y detected | Yes | B.1.351 |
|  | VoC-5 | 18-50yo; F | NP | 18.65 | N501Y detected | Yes | B.1.1.7 |
|  | VoC-6 | 18-50yo; M | NP | 21.38 | N501Y detected | Yes | B.1.1.7 |
|  | VoC-7 | >50yo; F | NP | 25.52 | N501Y detected | Yes | B.1.1.7 |
|  | VoC-8 | 18-50yo; F | SG | 34.02 | N501Y detected | Yes | B.1.1.7 |
|  | VoC-9 | 18-50yo; M | SG | 30.02 | N501Y detected | Yes | B.1.351 |
|  | VoC-10 | <18yo; F | NA | 16.94 | N501Y detected | Yes | B.1.1.7 |
|  | VoC-11 | >50yo; F | NA | 23.09 | N501Y detected | Yes | B.1.1.7 |
|  | VoC-12 | 18-50yo; M | SG | 23.46 | N501Y detected | Yes | B.1.1.7 |
|  | VoC-13 | <18yo; F | NP | 29.67 | N501Y detected | Yes | B.1.1.7 |
|  | VoC-14 | 18-50yo; M | SG | 26.97 | N501Y detected | Yes | B.1.1.7 |
|  | VoC-15 | 18-50yo; M | NP | 17.04 | N501Y detected | Yes | B.1.1.7 |
|  | VoC-16 | 18-50yo; F | NP | 24.88 | N501Y detected | Yes | B.1.1.7 |
|  | VoC-17 | <18yo; M | NA | 20.17 | N501Y detected | Yes | B.1.1.7 |
|  | VoC-18 | 18-50yo; M | SG | 26.83 | N501Y detected | Yes | B.1.1.7 |
|  | VoC-19 | <18yo; M | NP | 27.29 | N501Y detected | Yes | B.1.1.7 |
|  | VoC-20 | 18-50yo; M | NP | 22.54 | N501Y detected | Yes | B.1.1.7 |
|  | VoC-21 | 18-50yo; M | NA | 17.14 | N501Y detected | Yes | B.1.1.7 |
|  | VoC-22 | 18-50yo; M | NP | 28.19 | N501Y detected | Yes | B.1.1.7 |
| **Direct**<br>**WGS** | VoC-23 | 18-50yo; F | NP | NA | Background surveillance | Yes | B.1.1.7 |
|  | VoC-24 | <18yo; F | SG | 28.42 | Background surveillance | Yes | B.1.1.7 |
|  | VoC-25 | 18-50yo; M | NP | 27.70 | Background surveillance | Yes | B.1.351 |
|  | VoC-26 | 18-50yo; F | NP | 15.98 | Background surveillance | Yes | B.1.1.7 |
|  | VoC-27 | 18-50yo; F | NP | 34.83 | Cluster investigation | Yes | B.1.1.7 |
|  | VoC-28 | >50yo; M | SG | 25.96 | Targeted surveillance | Yes | B.1.1.7 |

Ct: cycle threshold; F: female; M: male; NA: not available; NP: nasopharyngeal; SG: saline gargle; VoC: variant of concern; WGS: whole genome sequencing; yo: years old.

## Discussion

Results from this point prevalence study of over 3,000 specimens demonstrated the feasibility of a province-wide combined VoC screening strategy incorporating WGS and N501Y qPCR screening, and showed low prevalence of VoCs with predominance of B.1.1.7 lineage in British Columbia in early 2021. This assessment provided the critical prevalence data required to benchmark VoC burden, and to inform planning of an optimized VoC detection strategy moving forward.

The BCCDC PHL currently performs the highest proportion of WGS in Canada, with up to 25% of SARS-CoV-2-positive specimens sequenced. However, this high-complexity approach necessitates specialized equipment and expertise, and computationally-intensive analysis that prolongs result turnaround time. Thus, even in specialized centralized settings, VoC testing alternatives are needed. In this study, given that most cases identified by N501Y screen were confirmed as VoCs, increasing the proportion of PCR-based screening over WGS upfront was deemed appropriate. Based on these data, the N501Y screening strategy was expanded to clinical laboratories across the province to decentralize testing, increase testing capacity, and reduce turnaround time to presumptive VoC identification. Subsequent point prevalence assessments based on this decentralized model are now planned for ongoing surveillance. However, a limitation of this approach is that emergence of variants devoid of N501Y mutation such as the recently-recognized B.1.525, reported here, may be underestimated due to failure of detection by qPCR targets employed, and requires alternative qPCR assays for identification (9, 10).

In summary, this study demonstrated the feasibility of a combined targeted and WGS testing strategy for VoC prevalence assessment in one Canadian province, which will be instrumental for continued VoC surveillance, and to inform public health efforts.

## Data Availability

Data are available from the corresponding author upon reasonable request.

## Acknowledgments

We thank the molecular, virology, bacteriology staff and microbiologists of the BCCDC Public Health Laboratory for their contribution toward testing, without which this work would not have been possible. We thank Rebecca Hickman, BSc and Ji-In Hum, BSc for their contributions toward test procedure development and result interpretation. We thank Chris Fjell, PhD, Braeden Klaver, MPH and Yayuk Joffres, MPH, and the BCCDC data analytics team for implementing the data collection and entry systems that enabled this work. We thank Drs. Nancy Matic, MD and Christopher Lowe, MD and the virology laboratory staff from St. Paul’s Hospital for sharing and verifying VoC data. We also thank the British Columbia Association of Medical Microbiologists for sharing samples and data that enabled province-wide data collection, and testing.

## Supplementary methods

### N501Y mutation screening

#### BCCDC

The British Columbia Centre for Disease Control (BCCDC) public health laboratory developed a real-time, duplex reverse transcription polymerase chain reaction assay targeting the envelope (*E*) gene and the N501Y mutation (A23063T) of the Spike (S) protein. The *E* gene probe was used to confirm detection of SARS-CoV-2 RNA in the specimen, as previously described by Corman *et al*.(11) The N501Y mutation is detected in the three main currently circulating Variants of Concern (VoCs): lineage B.1.1.7 (variant first described in the UK), B.1.351 (variant first described in South Africa), and lineage P.1 (variant described in Brazil). Briefly, total nucleic acids were extracted from 200μL of upper respiratory specimen matrix (universal transport medium or saline) using the Applied BioSystems MagMax™ Express 96 Nucleic Acid Extractor and the MagMax Viral/Pathogen Nucleic Acid Isolation Kit (Thermo Fisher Scientific, Waltham, MA) according to the manufacturer’s recommendations. Real-time RT-PCR was performed using the TaqMan Fast Virus Master Mix (Thermo Fisher Scientific) on the Applied Biosystems 7500 FAST real-time PCR system (Thermo Fisher Scientific). PCR set-up was performed using a volume of 5µL of patient specimen extract, for a final reaction volume of 20µL.

#### St. Paul’s Hospital

The St. Paul’s Hospital Clinical Microbiology Laboratory developed a real-time, reverse transcription polymerase chain reaction assay targeting the *E* gene and the N501Y mutation (A23063T) of the Spike (S) protein. Following extraction of viral RNA, SARS-CoV-2 detection was performed using the LightMix® ModularDx SARS-CoV (COVID19) E-gene assay (TIB Molbiol, Berlin, Germany), or with the combined extraction and amplification automated cobas® SARS-CoV-2 Test (Roche Molecular Diagnostics, Laval, QC) using the cobas® 6800 instrument. A smaller number of clinical samples were tested using Xpert® Xpress SARS-CoV-2/Flu/RSV (Cepheid, Sunnyvale, CA). Positives for SARS-CoV-2 were re-extracted and then screened for N501Y Spike (S) protein mutation with the VirSNiP SARS-CoV-2 Mutation Assays for Strain Surveillance (TIB Molbiol) using Roche LightCycler 480® (Roche Molecular Diagnostics, Laval, QC). For the PCR reaction, 5 µL of extracted sample were added to the reaction mixture as per the manufacturer’s instructions, and results were interpreted based on an analysis of amplification and melting curves.

#### BCCDC WGS testing

Whole genome sequencing was performed at the BCCDC PHL. In brief, SARS-CoV-2 RNA was extracted using the Applied BioSystems MagMax™ Express 96 Nucleic Acid Extractor and the MagMax Viral/Pathogen Nucleic Acid Isolation Kit (Thermo Fisher). Viral RNA was reverse transcribed into cDNA using Thermo SuperScript IV, and a two pool multiplex PCR with primers tiled across the entire SARS-CoV-2 genome, in 29 ∼1200bp segments, was performed.(12) DNA libraries for whole genome sequencing on a MiSeq instrument (Illumina, San Diego) were prepared using DNA Prep Library Preparation Kit (Illumina). SARS-CoV-2 whole genome consensus sequences and mutation compositions were generated using a modified Nextflow pipeline for running the ARTIC network’s (https://artic.network/ncov-2019) fieldbioinformatics tools (https://github.com/BCCDC-PHL/ncov2019-artic-nf). Reports detailing SARS-CoV-2 lineage information, sequencing QC metrics and mutational profiles were generated using ncov-tools from the Simpson Lab (https://github.com/jts/ncov-tools).

#### Statistical analysis

Exact 95% confidence intervals for proportions were calculated using Stata v.15.1 (Stata Corp, College Station, TX).

---

**Appendix Table 1.** Variant of concern N501Y screening post-point prevalence study period, British Columbia, Canada, February-March 2021

| **2021**<br>**Epiweek** | **Start date** | **Total**<br>**positive** | **Total**<br>**screened** | **Proportion**<br>**screened by**<br>**N501Y assay** | **Presumptive**<br>**VoC**<br>**positive** | **Presumptive**<br>**VoC positivity** |
| --- | --- | --- | --- | --- | --- | --- |
| 5 | 2021-01-31 | 3128 | 2125 | 67.93% | 23 | 1.08% |
| 6 | 2021-02-07 | 3116 | 1978 | 63.48% | 56 | 2.83% |
| 7 | 2021-02-14 | 3494 | 2442 | 69.89% | 108 | 4.42% |
| 8 | 2021-02-21 | 3653 | 3064 | 83.88% | 231 | 7.54% |
| 9 | 2021-02-28 | 3931 | 3557 | 90.49% | 434 | 12.20% |

VoC: variant of concern.

**Appendix Figure 1.**
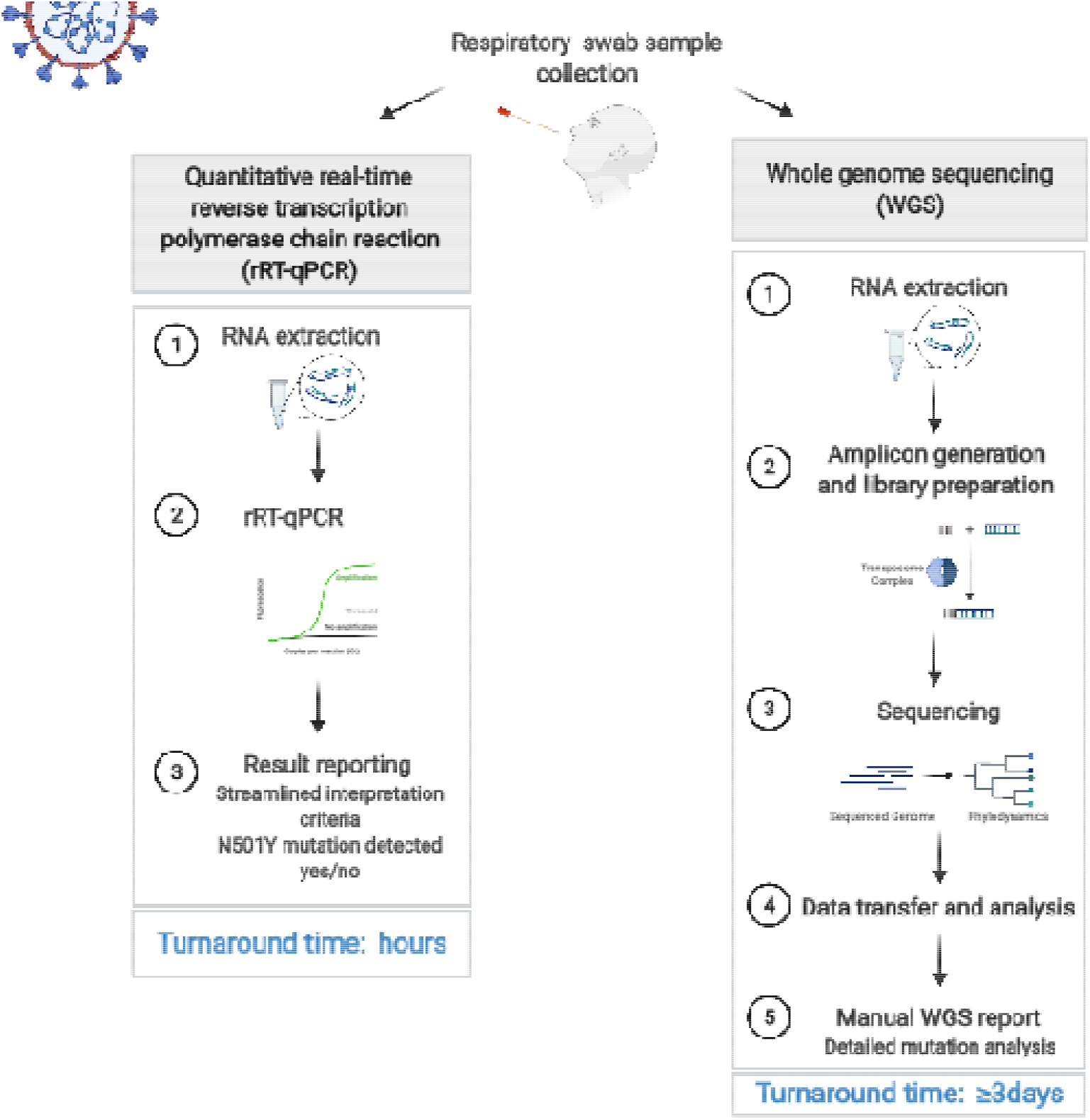
Comparative analytical workflow of real-time reverse transcription polymerase chain reaction testing for N501Y mutation detection (left) vs whole genome sequencing (WGS) (right). The WGS process is significantly more complex, requires additional expertise and longer turnaround time than conventional PCR. A VoC detection strategy that leverages the uses of both approaches is needed to optimize laboratory testing capacity and turnaround time. (Figure created with BioRender) RNA: ribonucleic acid; rRT-PCR: real-time reverse transcription polymerase chain reaction; WGS: whole genome sequencing.

**Appendix Figure 2.**
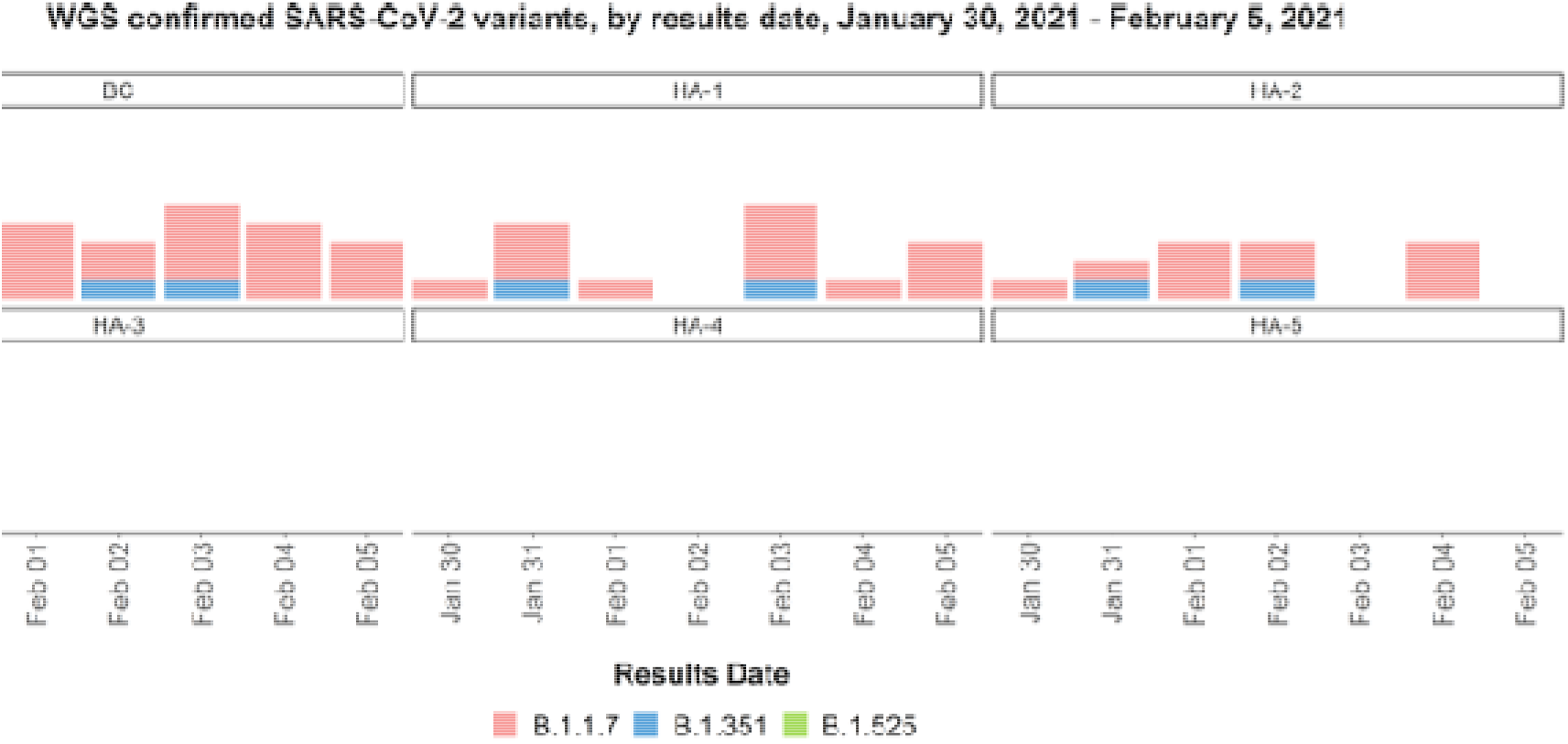
Number of confirmed variants of concern (VoCs) in British Columbia, from January 30^th^ to February 5^th^ 2021 by resulting date. Geographical stratification of the data revealed that the confirmed VOC cases were concentrated in two health authorities, enabling focused public health efforts. Lineage B.1.1.7 strongly predominated, there was a single B.1.525 case, and there was no lineage P.1 identified during this time period. BC: British Columbia; HA: health authority; Feb: February; SARS-CoV-2: severe acute respiratory syndrome coronavirus 2; WGS: whole genome sequencing.

## References

1. Walensky RP, Walke HT, Fauci AS. 2021. SARS-CoV-2 Variants of Concern in the United States—Challenges and Opportunities. JAMA doi:10.1001/jama.2021.2294.

2. Galloway SE, Paul P, MacCannell DR, Johansson MA, Brooks JT, MacNeil A, Slayton RB, Tong S, Silk BJ, Armstrong GL, Biggerstaff M, Dugan VG. 2021. Emergence of SARS-CoV-2 B.1.1.7 Lineage - United States, December 29, 2020-January 12, 2021. MMWR Morb Mortal Wkly Rep 70:95–99.

3. Muik A, Wallisch AK, Sanger B, Swanson KA, Muhl J, Chen W, Cai H, Maurus D, Sarkar R, Tureci O, Dormitzer PR, Sahin U. 2021. Neutralization of SARS-CoV-2 lineage B.1.1.7 pseudovirus by BNT162b2 vaccine-elicited human sera. Science 371:1152–1153.

4. Davies NG, Abbott S, Barnard RC, Jarvis CI, Kucharski AJ, Munday JD, Pearson CAB, Russell TW, Tully DC, Washburne AD, Wenseleers T, Gimma A, Waites W, Wong KLM, van Zandvoort K, Silverman JD, Group CC-W, Consortium C-GU, Diaz-Ordaz K, Keogh R, Eggo RM, Funk S, Jit M, Atkins KE, Edmunds WJ. 2021. Estimated transmissibility and impact of SARS-CoV-2 lineage B.1.1.7 in England. Science doi:10.1126/science.abg3055.

5. Challen R, Brooks-Pollock E, Read JM, Dyson L, Tsaneva-Atanasova K, Danon L. 2021. Risk of mortality in patients infected with SARS-CoV-2 variant of concern 202012/1: matched cohort study. BMJ 372:579.

6. GISAID. 2021. Tracking of Variants. https://www.gisaid.org/hcov19-variants/. Accessed February 20 2021.

7. ECCDC. 2021. Sequencing of SARS-CoV-2: first update.

8. ECCDC. 2021. Methods for the detection and identification of SARS-CoV-2 variants. https://www.ecdc.europa.eu/sites/default/files/documents/Methods-for-the-detection-and-identification-of-SARS-CoV-2-variants.pdf. Accessed March 13 2021.

9. GISAID. 2021. B.1.525 2021-02-20. https://cov-lineages.org/global_report_B.1.525.html. Accessed March 12 2021.

10. Bal A, Destras G, Gaymard A, Stefic K, Marlet J, Eymieux S, Regue H, Semanas Q, d’Aubarede C, Billaud G, Laurent F, Gonzalez C, Mekki Y, Valette M, Bouscambert M, Gaudy-Graffin C, Lina B, Morfin F, Josset L, Group CO-DHS. 2021. Two-step strategy for the identification of SARS-CoV-2 variant of concern 202012/01 and other variants with spike deletion H69-V70, France, August to December 2020. Euro Surveill 26.

11. Corman VM, Landt O, Kaiser M, Molenkamp R, Meijer A, Chu DKW, Bleicker T, Brunink S, Schneider J, Schmidt ML, Mulders D, Haagmans BL, van der Veer B, van den Brink S, Wijsman L, Goderski G, Romette JL, Ellis J, Zambon M, Peiris M, Goossens H, Reusken C, Koopmans MPG, Drosten C. 2020. Detection of 2019 novel coronavirus (2019-nCoV) by real-time RT-PCR. Euro Surveill 25:1–8.

12. Freed NE, Vlkova M, Faisal MB, Silander OK. 2020. Rapid and inexpensive whole-genome sequencing of SARS-CoV-2 using 1200 bp tiled amplicons and Oxford Nanopore Rapid Barcoding. Biol Methods Protoc 5:bpaa014.

## References

1. Corman VM, Landt O, Kaiser M, et al. Detection of 2019 novel coronavirus (2019-nCoV) by real-time RT-PCR. Euro Surveill.2020;25(3):1–8.

2. Freed NE, Vlkova M, Faisal MB, Silander OK. Rapid and inexpensive whole-genome sequencing of SARS-CoV-2 using 1200 bp tiled amplicons and Oxford Nanopore Rapid Barcoding. Biol Methods Protoc.2020;5(1):bpaa014.

